# Rural-Urban Disparities in Near-Completed Fertility among Women Aged 40-49 in Nigeria: Evidence from the 2024 Nigeria Demographic and Health Survey

**DOI:** 10.64898/2026.07.28.26359080

**Authors:** Olamide Martins Ajayi, Olubusayo Bankole Ogunsemoyin, Aboluwaji Daniel Ayinmoro

## Abstract

Rural-urban fertility differences may reflect unequal distributions of education, household resources, union timing, and regional characteristics rather than an independent effect of residence. This study examined whether the rural-urban gap in near-completed fertility among Nigerian women aged 40-49 persisted after accounting for these compositional factors. Data were drawn from 7,370 women in the 2024 Nigeria Demographic and Health Survey. Children ever born were analyzed using weighted descriptive statistics, residence-specific comparisons, and four survey-weighted Poisson generalized linear models with a log link and cluster-robust standard errors. The weighted mean number of children ever born was 5.53, ranging from 4.68 among urban women to 6.39 among rural women. In the age-adjusted model, rural women had 37% more children than urban women (IRR = 1.37, p < .001), but residence was no longer statistically significant after adjustment for education and household wealth. In the fully adjusted model, women aged 45-49 had more children than those aged 40-44 (IRR = 1.09, p < .001). Secondary and higher education were associated with lower fertility (IRR = 0.92 and 0.82, respectively; p < .001), as was residence in the richest households (IRR = 0.84, p < .001). First cohabitation at ages 20-24 and 25 or older was also associated with lower fertility (IRR = 0.82 and 0.64; p < .001). The rural-urban fertility gap largely reflected socioeconomic, marital-timing, and regional inequalities.

## Introduction

Fertility remains a key factor in Nigeria’s demographic change, population policies, and long-term development strategies. The country is experiencing a regional fertility transition that is progressing unevenly. Although the global demographic shift is generally described as moving from high mortality and fertility to longer lives and smaller families, its timing and social distribution vary significantly across countries and within populations (Hirschman, 1994; United Nations DESA, Population Division, 2024). In sub-Saharan Africa, this transition has been slower and more uneven internally compared to other regions. Nigeria exemplifies this diversity, with fertility rates varying by residence, region, education, religion, household wealth, and marriage timing (Bongaarts & Casterline, 2013; Bongaarts, 2017; NPC & ICF, 2019). Analyzing rural versus urban residence is particularly helpful because it reflects disparities in life chances. Rural and urban women face different opportunities in schooling, labor markets, transportation, healthcare, legal protections, media access, and social norms regarding marriage and childbearing. These factors accumulate over a woman’s life and collectively influence fertility. Rural living may sustain higher fertility due to early unions, limited education, fewer material resources, stronger kin-based reproductive oversight, and less access to family planning. Conversely, urban residence may reduce fertility by providing greater education, more employment opportunities, higher child-rearing costs, and greater access to reproductive health information. The key question is not just why rural women have more children, but whether rural residence has a significant impact after accounting for socioeconomic and life-course differences across rural and urban populations.

This article focuses on women aged 40-49, a critical age group because they represent women whose reproductive histories are mostly, though not entirely, complete. Gathering data on children ever born among women in this later reproductive phase provides a cumulative fertility measure that is less influenced by short-term timing fluctuations than period fertility indicators. Nonetheless, since women aged 40-44 might still have additional children, the term “near-completed fertility” is used instead of “strict completed fertility.” This distinction is important; it avoids overestimating fertility and aligns with the 2024 Nigeria Demographic and Health Survey data, which includes women aged 15-49 and allows analysis of the 40-49 age group (Federal Ministry of Health and Social Welfare of Nigeria [FMoHSW], NPC, & ICF, 2025; Croft et al., 2023). Period fertility indicators, like the total fertility rate, remain valuable but measure fertility over a specific timeframe rather than the lifetime reproductive experience of a cohort. Conversely, children ever born reflect the total outcome of past marriage patterns, contraceptive use, childbearing preferences, social norms, and economic factors. For women aged 40-49, this measure is practical for examining how structural inequalities influence realized fertility over the reproductive lifespan (Bongaarts, 1978). In Nigeria, where fertility histories are affected by disparities in education, early cohabitation, and regional differences, this comprehensive approach is particularly pertinent.

The study has three primary objectives. First, it assesses the rural-urban gap in near-completed fertility among women aged 40-49 using 2024 NDHS data. Second, it compares rural and urban women based on education, household wealth, geopolitical zone, religion, union timing, employment, and contraceptive use. Third, it investigates whether this crude fertility difference remains after adjusting for these factors. This approach treats residence not as a direct cause but as a social context influenced by or confounded with compositional differences. The article brings three contributions to demographic research: recent data from the 2024 NDHS, a focus on women aged 40-49 to emphasize cumulative fertility rather than current fertility, and a connection of the fertility gap to theories on demographic transition, wealth flows, proximate determinants, diffusion, gender, and education. This is crucial because high fertility in rural Nigeria cannot be explained solely by contraceptive access; it is rooted in broader social, economic, and cultural structures (Caldwell, 1976, 1982; Cleland et al., 2006; McDonald, 2000). Analyzing women aged 40-49 allows the study to link cohort effects with current policies. Births among these women represent not only current conditions but also influences from adolescence to adulthood, such as school systems, marriage norms, contraceptive environments, and household economies. The rural-urban differences at this age reflect accumulated inequality over time rather than current residence alone. This distinction is especially important in Nigeria, where a woman’s place at survey time may symbolize years of unequal access to education and reproductive health, despite the cross-sectional survey. Nigeria’s demographic diversity means fertility varies widely across residence, region, education, and household resources. A national fertility estimate conceals these social disparities where high fertility persists. For policy, the key issue is not only declining fertility but also understanding which groups are affected by social and institutional factors that facilitate this decline. The 2024 NDHS provides recent, representative data and variables to explore rural-urban differences alongside education, wealth, union timing, and religion (FMoHSW et al., 2025). Focusing on women aged 40-49 emphasizes the life-course aspect of fertility. By this age, most women have completed most of their childbearing, although some births may still occur before age 50. The number of children ever born in this group reflects the cumulative effects of early-life factors, such as childhood environment, education, marriage timing, household economic status, and access to reproductive health information. This differs from analyzing only recent births, which may capture short-term fluctuations rather than the full reproductive history of women nearing the end of their reproductive years. The article avoids two common pitfalls: first, treating rural residence as an cultural explanation for high fertility, which can reinforce stereotypes and overlook institutional influences; second, viewing fertility only through contraceptive use, neglecting how schooling, gender roles, marriage patterns, family size ideals, and service quality shape contraceptive behavior (Bongaarts, 1978; Cleland et al., 2006; Tsui et al., 2017).

For a comprehensive rural-urban analysis, it is essential to understand the social mechanisms linking residence and cumulative births. Nigeria’s data also require careful interpretation, as national averages may obscure significant differences across geopolitical zones, education levels, and household wealth. Previous Nigeria DHS studies revealed substantial variations in fertility timing and levels across six zones, with education playing a key role (Olowolafe et al., 2023, 2025). The 2024 NDHS enables us to revisit these issues by examining whether residential disparities among older women persist after accounting for compositional factors. The analytical payoff is policy-relevant. If rural residence remains strongly associated with near-completed fertility after adjustment, then place-targeted interventions may need to address the unmeasured rural context more directly, including community norms, service availability, and local marriage systems. If the rural effect weakens after adjusting for education, wealth, and cohabitation timing, then the residence gap is mainly the demographic consequence of unequal social composition. The present study shows the second pattern. That finding does not make rural context irrelevant; rather, it specifies where policy leverage is likely to be strongest.

## Background

### Rural-Urban Fertility Differentials

Sub-Saharan Africa has become the focal point in current discussions about fertility transition due to its persistently high fertility rates and uneven decline across different countries and social groups. Bongaarts and Casterline (2013) note that Africa’s fertility transition is unique because factors like desired family size, contraceptive adoption, postpartum practices, and socioeconomic conditions follow distinctive patterns. Bongaarts (2017) adds that the transition in Africa is not missing but rather delayed, uneven, and sometimes halted. Shapiro and Gebreselassie (2008) describe a similar pattern characterized by decline interrupted by periods of stagnation, while Ezeh et al. (2009) observe that fertility stalls can occur when progress in education, contraceptive use, and reproductive preferences is insufficient to sustain decline.

Nigeria aligns with this continental trend. Previous NDHS reports and studies using Nigerian DHS data indicate that fertility is unevenly distributed geographically, with the North-West and North-East regions typically showing higher fertility rates than southern areas. Rural regions also consistently report higher fertility than urban ones (NPC & ICF, 2019; Olowolafe et al., 2023). The 2024 NDHS offers a new opportunity to re-examine these patterns, taking into account more recent demographic trends. It is especially valuable because it allows analysis of women aged 40-49, whose total number of children reflects the long-term reproductive history accumulated over decades (FMoHSW et al., 2025; ICF, 2025).

The rural-urban fertility gap remains a consistent feature of demographic transition. Urban areas tend to see fertility decline sooner because they have concentrated access to schools, healthcare, salaried employment, public administration, transportation, mass media, and evolving family norms. In contrast, rural areas often experience later declines due to stronger influences from household labor systems, kinship obligations, farming practices, limited education, and early marriages. Studies across Africa, Asia, and Latin America show that female education significantly reduces rural-urban differences in fertility. However, residence interacts with socioeconomic factors, rather than acting independently (Adhikari et al., 2025; Ainsworth et al., 1996).

### Education, Marriage Timing, and Socioeconomic Stratification

A rural-urban differential is rarely only a matter of physical location. Residence is a spatial marker of schooling systems, labor markets, infrastructure, health-service density, child-survival contexts, and exposure to demographic innovations. In many African settings, fertility decline has tended to begin in urban areas and among women with more schooling before diffusing more unevenly to rural populations (Bongaarts & Casterline, 2013; Shapiro & Gebreselassie, 2008). This does not mean that rural women are culturally fixed in high-fertility regimes. It means that the conditions that support later marriage, smaller desired families, and effective contraception often arrive later, less consistently, or in less usable forms in rural communities.

The Nigerian case is especially important because the rural-urban contrast intersects with the geopolitical region. Northern and southern zones differ in schooling, union formation, religion, and socioeconomic conditions, while rural residence is more common in some high-fertility zones. Treating residence as a single explanatory variable without considering these overlaps risks attributing to rural culture what may instead reflect the concentration of disadvantage. Conversely, ignoring residence would miss the spatial organization of inequality that shapes the everyday conditions under which reproductive decisions are made.

Education is the most consistent socioeconomic correlate of fertility. Schooling can delay union formation, increase literacy, improve contact with health information, raise the opportunity cost of early and high-parity childbearing, and strengthen women’s capacity to negotiate reproductive preferences. Ainsworth et al. (1996), using DHS data across 14 sub-Saharan African countries, showed that women’s schooling was associated with both lower cumulative fertility and higher contraceptive use. Kravdal (2002) extended this argument by showing that community-level education also matters, suggesting that schooling influences fertility not only through individual attributes but also through the local normative and informational environment.

In Nigeria, education is also deeply spatialized. Rural women are more likely to have no formal education and less likely to reach secondary or higher education. This matters because the fertility consequences of low schooling are cumulative. Girls who leave school early are more likely to enter union earlier, begin childbearing earlier, and experience a longer reproductive exposure window. Conversely, secondary and higher education can delay marriage and first birth, increase aspirations beyond early motherhood, and create a social context in which a smaller family size becomes more plausible. Recent Nigeria-focused evidence confirms that education remains one of the strongest correlates of regional and individual fertility differences (Olowolafe et al., 2025).

Age at first cohabitation is a proximate life-course pathway through which structural inequalities become reproductive outcomes. In settings where childbearing remains strongly tied to marriage or co-residential union, earlier union entry extends the period during which births can occur. It also places women into reproductive decision-making contexts at younger ages, when schooling, labor-force entry, and bargaining power may be limited. Casterline et al. (2017) show that changes in age at marriage are closely linked to the onset and progression of fertility transition in sub-Saharan Africa. In Nigeria, rural residence, low schooling, and poverty often reinforce early union formation, making age at first cohabitation a key pathway linking residence to cumulative fertility.

Household wealth is another major axis of fertility stratification, but its interpretation requires caution. Wealth does not affect fertility only through income. It also captures living standards, housing conditions, access to transport, child-rearing costs, schooling opportunities, and access to health services. In agrarian or kinship-based settings, children may be valued for labor, lineage continuity, and old-age security. In more urbanized settings, the direct and indirect costs of education, housing, and health care may increase pressure toward smaller families. Caldwell’s wealth flows theory is useful here because it argues that high fertility remains rational where net economic and social flows move from children to parents. However, fertility declines when flows reverse, and parents make heavier investments in child quality (Caldwell, 1976, 1982).

### Religion, Region, and Normative Contexts

Education influences fertility indirectly by impacting the sequence of life events. It can postpone first cohabitation, boost chances of paid work, increase exposure to print and electronic media, and alter perceptions of the benefits of having fewer children (Ainsworth et al., 1996; Jejeebhoy, 1995). These effects accumulate over time. A woman who stays in school longer may start a union later, have children later, gain more bargaining power within marriage, and face higher opportunity costs from multiple births. For women aged 40-49, the total number of children ever born reflects educational experiences throughout their lives, rather than just their reproductive preferences at the time of the survey.

Wealth functions in a related yet distinct manner. DHS wealth quintiles are based on assets and serve as indicators of relative household living standards rather than direct income measures. In fertility studies, wealth reflects access to infrastructure, housing quality, consumer goods, education opportunities, and health services. It can also represent different household economies: in farming areas, children may continue to contribute economically and socially, whereas urban households might favor smaller families due to higher costs of housing, schooling, transportation, and healthcare. The wealth gradient in cumulative fertility thus encompasses both material and normative aspects. Additionally, religion and ethnicity are significant in Nigeria because they are interconnected with region, marriage customs, gender roles, and family-size ideals. Religious affiliation is not solely about belief; it can also indicate institutional authority, marriage practices, acceptance of contraception, and community expectations. Ethnicity similarly reflects histories of kinship, lineage, marriage, and social organization. Because religion, ethnicity, and region overlap considerably in Nigeria, treating each as a separate causal factor would be misleading. A robust empirical approach considers these variables as social-location controls, focusing mainly on education, wealth, and union timing, which are clearer mechanisms (Caldwell et al., 1992; McDonald, 2000).

### Ideational Diffusion and Gender Context

The literature on fertility transition highlights the importance of ideational diffusion. Cleland and Wilson (1987) challenged explanations that focus solely on economic factors, suggesting that fertility decline can also result from the spread of new ideas about family planning, modernity, and reproductive control. Similarly, Casterline (2001) emphasizes diffusion mechanisms like social learning and influence. These processes are particularly relevant to rural-urban differences, as urban areas often facilitate faster dissemination of reproductive ideas through schools, media, workplaces, peer groups, and health services. While rural areas are not completely isolated from information, the speed and trustworthiness of diffusion may vary where local authorities, kinship elders, and religious leaders dominate reproductive decision-making.

Gender theory enhances these perspectives. McDonald (2000) suggests that fertility outcomes depend in part on how gender equity is structured within families and institutions. When women gain educational opportunities, but family systems remain unequal, reproductive transitions may be delayed or inconsistent. Jejeebhoy (1995) also shows that women’s education, autonomy, and reproductive behavior are closely linked, though the relationship varies across contexts. This is especially relevant for Nigeria, where rural-urban differences also reflect gendered disparities in life-course power. The ability to delay marriage, stay in school, negotiate contraception, or limit births is not equally distributed. While contraceptive practice explains part of this, it should not be seen as the sole factor. Bongaarts’ proximate determinants framework identifies contraception as a key mechanism through which social factors influence fertility, alongside marriage, postpartum infecundability, abortion, and sterility (Bongaarts, 1978). Access to family planning and contraceptive demand are thus vital to fertility decline. Cleland et al. (2006) describe family planning as an unfinished global agenda, and Liu and Raftery (2020) demonstrate that education and family planning together can accelerate fertility decline. However, contraceptive use measured at ages 40-49 may reflect behaviors after most births have already occurred. Therefore, contraceptive methods reported at interview should be seen as a contemporaneous indicator of reproductive behavior, not a full history of contraceptive exposure.

Evidence from sub-Saharan Africa underscores this cautious approach. Tsui et al. (2017) reveal that contraceptive use varies by factors such as age, marital status, parity, policy settings, and the availability of services. Rossier and Corker (2017) highlight that traditional methods still play a significant role in some areas and should not be considered as non-use. In Nigeria, Mercer et al. (2019) find notable differences in family planning indicators across various states and age-parity groups. These results suggest that contraceptive use among women aged 40-49 may often stem from reasons like stopping, high parity, health concerns, or postpartum factors, rather than early fertility limitations.

### Theoretical Framework

Religion and ethnicity should not be viewed solely as doctrines or identities. Their demographic influence typically operates through local marriage customs, gender dynamics, kinship expectations, family size ideals, and the acceptability of contraception within communities. Nigerian fertility studies must, therefore, regard religion and ethnicity as socially embedded factors rather than mere attitudes. This explains why residence, religion, and region are analyzed together: each reflects a broader social context influencing fertility behavior. The ideational perspective is useful because fertility decline often depends on more than just access to services; it also involves spreading new norms about family size, the acceptability of fertility regulation, and the perceived compatibility of contraception with marital, religious, and gender norms (Casterline, 2001; Cleland & Wilson, 1987). Although media exposure was not included as a covariate in the final data model, it remains significant: information environments shape whether women and couples view smaller families as desirable, legitimate, and achievable.

This article’s theoretical position is therefore deliberately layered. Caldwell’s wealth flows theory explains why high fertility may remain socially meaningful in settings where children contribute labor, lineage security, and old-age support (Caldwell, 1976, 1982). Bongaarts’ proximate determinants framework clarifies how broader social conditions translate into births through exposure to union, contraception, and biological mechanisms (Bongaarts, 1978). Diffusion theory explains why fertility norms may change unevenly across rural and urban settings (Casterline, 2001; Cleland & Wilson, 1987). Gender and education perspectives explain why fertility decline depends on women’s life-course opportunities and bargaining power (Jejeebhoy, 1995; McDonald, 2000). Together, these perspectives justify an empirical model that asks whether rural residence remains associated with near-completed fertility after adjustment for education, wealth, and age at first cohabitation.

### Conceptual Expectations

Caldwell’s wealth flow theory is especially useful for understanding rural-urban differences in fertility because it connects fertility to the direction of intergenerational transfers (Caldwell, 1976, 1982). In contexts where children provide labor, social status, security, and help maintain family lineage, high fertility remains socially valuable. Conversely, when parents need to invest heavily in education, housing, health, and consumption, the economic and social rationale for smaller families becomes more compelling. Thus, differences between rural and urban areas in Nigeria can partly be seen as variations in household economies and expected returns from children. Bongaarts’ proximate determinants framework complements this by outlining the immediate social mechanisms influencing fertility (Bongaarts, 1978, 2017). In this article, age at first cohabitation is especially significant since it affects the duration of exposure to marital childbearing. While the contraceptive method also matters, as it is measured at the survey date, it mainly reflects current reproductive status rather than lifetime exposure. This distinction is particularly important for women aged 40-49, whose children ever born usually precede their current contraceptive use.

The conceptual expectation is clear: rural women aged 40-49 are expected to report a higher average number of children ever born compared to urban women. However, once education and household wealth are included in the analysis, the rural coefficient should decrease, as rural women typically lag in these factors. It is also expected to decline further, even after accounting for age at first cohabitation, since early union increases reproductive exposure. If a significant rural effect remains after these controls, it might indicate the influence of unmeasured community norms, service environments, or local marriage systems. Conversely, if the rural effect diminishes substantially, the rural-urban difference is likely due to socioeconomic and life-course factors rather than residence alone. The distinction between period fertility and cumulative fertility is also conceptually important: the total fertility rate estimates the number of births a hypothetical woman would have if current fertility rates applied throughout her reproductive life, useful for analyzing current conditions but sensitive to timing shifts. Children ever born among women aged 40-49 reflect the actual accumulated live births by late reproductive age, representing a near-cohort’s reproductive history. The DHS system allows for both analyses, but each indicator should be interpreted based on what it measures (Croft et al., 2023).

Using children ever born shifts focus from immediate behavior to the buildup of structural exposure. A woman with many children by age 45 did not develop that fertility level in a single year but through earlier life events such as leaving school early, first cohabitation, birth intervals, partner choices, contraceptive use, child survival, household labor demands, and social expectations. This makes cumulative fertility valuable for examining inequality, as it reflects the long-term impacts of social position. Especially in rural-urban studies, it captures not just current service access but also the social institutions shaping women’s life courses. The educational gradient in fertility is not solely about contraception knowledge; education influences marriage timing, the meaning of adulthood, the opportunity costs of early motherhood, health literacy, exposure to networks, bargaining power, and acceptance of smaller families. While aligning with classical demographic transition theory, these mechanisms also highlight gender roles and agency (Jejeebhoy, 1995; McDonald, 2000). In rural Nigeria, where school access is uneven, education likely acts as both a direct and indirect route to lower cumulative fertility.

The wealth gradient should be understood as more than just household purchasing power. In DHS analyses, wealth quintiles are asset-based, relative measures that summarize dwelling conditions, ownership of durable goods, and access to utilities. They do not equate to income but reflect material status within the national context. This is useful for fertility studies because household assets correlate with access to education, child-rearing costs, transportation, media exposure, and health services. When the wealthiest women have fewer children than the poorest, it likely indicates multiple advantages rather than a single economic factor. The timing of marriage and cohabitation clearly links social structure to fertility, especially in Nigeria and much of sub-Saharan Africa, where childbearing remains closely tied to union formation. Early cohabitation increases pregnancy risk and limits time for schooling and labor-market participation before motherhood. It may also place women in households where reproductive choices are influenced by husbands, senior relatives, and community norms. Casterline et al. (2017) highlight that changes in marriage timing are strongly associated with the start of fertility transition in sub-Saharan Africa. This analysis applies that insight to Nigeria’s near-completion of fertility decline.

The Nigerian data also highlight the importance of considering the intersection of religion, region, and residence. The North-West and North-East differ significantly from southern areas in terms of fertility, education history, marriage age, poverty, and religious makeup. Viewing religion solely as an independent factor would be insufficient analytically. Likewise, treating geopolitical zones merely as control variables may underestimate their role as carriers of distinct institutional and historical characteristics. The comprehensive model includes these variables to prevent overstating the impact of residence, but they are primarily seen as indicators of broader social context rather than direct causal factors. Evidence from family planning in Nigeria supports this cautious approach. Mercer et al. (2019) find that contraceptive use, unmet need, and demand satisfaction vary widely across states and demographic groups, showing that national indicators mask regional heterogeneity. This logic also applies to fertility. Nigeria’s rural areas are diverse, and even urban regions show significant inequality. Nonetheless, comparing rural versus urban areas nationally remains insightful, as it highlights a fundamental social divide and indicates whether this divide is mostly contextual or persists after adjustment for compositional differences.

The literature on fertility stalls offers further context. According to Shapiro and Gebreselassie (2008) and Ezeh et al. (2009), fertility decline may slow or halt when improvements in education, contraceptive use, child survival, and desired family size do not reinforce each other. Nigeria’s rural-urban pattern should be interpreted within this framework. The continued high fertility rates among rural women aged 40-49 do not necessarily indicate an absence of decline. Instead, it suggests that the cumulative conditions fostering decline were weaker for women whose reproductive years occurred amid lower levels of education, earlier unions, and fewer household resources. This background yields a practical expectation: the rural-urban gap should be prominent in descriptive analyses but should narrow in multivariable models once factors such as education, wealth, and initial cohabitation are accounted for. If the coefficient diminishes or becomes nonsignificant, it does not mean rural life is demographically irrelevant. Rather, it implies that observable socioeconomic and life-course variables account for much of the observed rural disadvantage. This distinction is critical for policy, as it shifts the focus from merely providing services in rural areas to addressing the conditions shaping rural women’s reproductive lives.

## Data and Methods

### Data Source and Study Population

This study utilized the women’s individual recode file from the 2024 Nigeria Demographic and Health Survey (NDHS). The NDHS is a nationally representative, cross-sectional household survey conducted by Nigeria’s Federal Ministry of Health and Social Welfare and the National Population Commission, with technical support from ICF via The DHS Program (Federal Ministry of Health and Social Welfare of Nigeria [FMoHSW], National Population Commission [NPC], & ICF, 2025). It employed a stratified, two-stage cluster sampling design and collected standardized data on fertility, union formation, contraceptive use, employment, and socioeconomic factors. The study population included all women aged 40-49 who participated in the survey and were included in the provider dataset (unweighted n = 7,370). The age criterion was set before analysis to focus on women nearing the end of their reproductive years. For women aged 40-49, the number of children ever born reflects near-completed fertility, as some women, especially those aged 40-44, may have additional children later in life. Access to the 2024 NDHS datasets was requested via The DHS Program on May 3, 2026, with ICF granting access on May 4, 2026.

### Outcome and Explanatory Variables

The outcome was the total number of children ever born, measured using the DHS variable V201. This variable records the cumulative number of live births reported by each woman at interview and is a non-negative count. A secondary categorization was employed to distinguish women with fewer than five children from those with five or more, solely to illustrate high-parity childbearing in Table 2; it was not used as a regression outcome.

The main explanatory variable was place of residence (V102), classified as urban or rural, with urban as the reference category. Age (V013) was categorized as 40-44 or 45-49 years. Education (V106) was grouped into no education, primary, secondary, and higher levels. Household wealth (V190) was divided into the poorest, poorer, middle, richer, and richest. The geopolitical zone (V101) included North-Central, North-East, North-West, South-East, South-South, and South-West. Religion (V130) was recoded into Christianity, Islam, and traditional/other religions. Current employment status (V714) was recorded as ‘no’ or ‘yes’. Age at first cohabitation (V511) was classified as before age 18, 18-19, 20-24, 25 or older, or never in union/not stated. The final combined category included women with no available age at first cohabitation, either because they never entered a union or the age was not specified. The current contraceptive method (V312) was reclassified into not using, traditional methods, or modern methods. Current union status (V502) was included in the descriptive profile but not in the count models alongside age at first cohabitation, as these variables overlap in measuring union exposure, and age at first cohabitation is the preferred life-course measure.

### Statistical Analysis

The analysis was conducted in four stages. First, weighted percentages outlined the sociodemographic, union, employment, and contraceptive profiles of women aged 40-49. Second, the weighted mean number of children ever born and the percentage of women with five or more children were calculated across different characteristics. Third, residence-specific column percentages for education, household wealth, and age at first cohabitation were determined to highlight the main compositional differences between rural and urban populations. Fourth, sequential Poisson generalized linear models with a log link estimated the relationships with the expected number of children ever born. Poisson regression was chosen because the outcome is a count, and incidence rate ratios offer a straightforward multiplicative interpretation of differences in cumulative births (Cameron & Trivedi, 2013). The full model had a residual deviance-to-degrees-of-freedom ratio of about 0.98, and robust sandwich standard errors were applied to account for potential variance misspecification and within-cluster dependence.

Model 1 considered residence and age group. Model 2 incorporated education and household wealth to evaluate how much the crude residence coefficient reflected socioeconomic factors. Model 3 included age at first cohabitation to measure the length of union-based exposure to childbearing. Model 4 further controlled for religion, geopolitical zone, current employment, and contraceptive method. This sequence was based on theory and aimed to illustrate how the residence coefficient evolved with additional socioeconomic and life-course variables; it was not intended as a formal causal mediation analysis.

All estimates used the women’s individual sampling weight (V005/1,000,000). Regression standard errors were computed with a cluster-robust sandwich estimator at the primary sampling unit level (V021). Coefficients were exponentiated and reported as incidence rate ratios (IRRs) with 95% confidence intervals in Table 4. Statistical tests were two-sided; p < .05 was considered significant. The reference groups included urban residence, ages 40-44, no education, the lowest wealth quintile, first cohabitation before age 18, Christianity, North-Central residence, not currently employed, and no contraceptive use. Analyses were conducted in SPSS v29.

### Ethical Considerations, Consent, and Data Access

This study conducted a secondary analysis using de-identified public survey data, involving no direct contact with respondents. The original NDHS protocol was reviewed for ethical approval by the relevant Nigerian authorities and the ICF Institutional Review Board. Before interviews, survey implementers obtained informed consent from participants (FMoHSW et al., 2025). The dataset contained no personal identifiers such as names, addresses, images, or biomaterials, eliminating the need for additional consent for this secondary analysis. Use of the data adhered to The DHS Program’s data-access policies. A request to access the 2024 NDHS datasets was submitted on May 3, 2026, and approved by ICF on May 4, 2026. The data remains accessible to registered researchers through The DHS Program upon approval of a research proposal (ICF, 2025).

## Results

### Weighted Sample Profile

Table 1 presents the weighted sociodemographic and reproductive profile of women aged 40–49 years included in the 2024 Nigeria Demographic and Health Survey. Most respondents were aged 40–44 years (57.3%), while 42.7% were aged 45–49 years. The women were almost evenly distributed between urban (50.6%) and rural (49.4%) areas. Educational attainment was generally low: 40.5% had no formal education, and 17.3% had only primary education. Slightly more than one-quarter had secondary education (27.0%), while 15.3% had attained higher education. The distribution across household wealth categories showed a gradual increase from the poorest to the richest households, with the largest proportion of women in the richest wealth quintile (26.7%) and the smallest in the poorest quintile (16.9%).

**Table 1.**
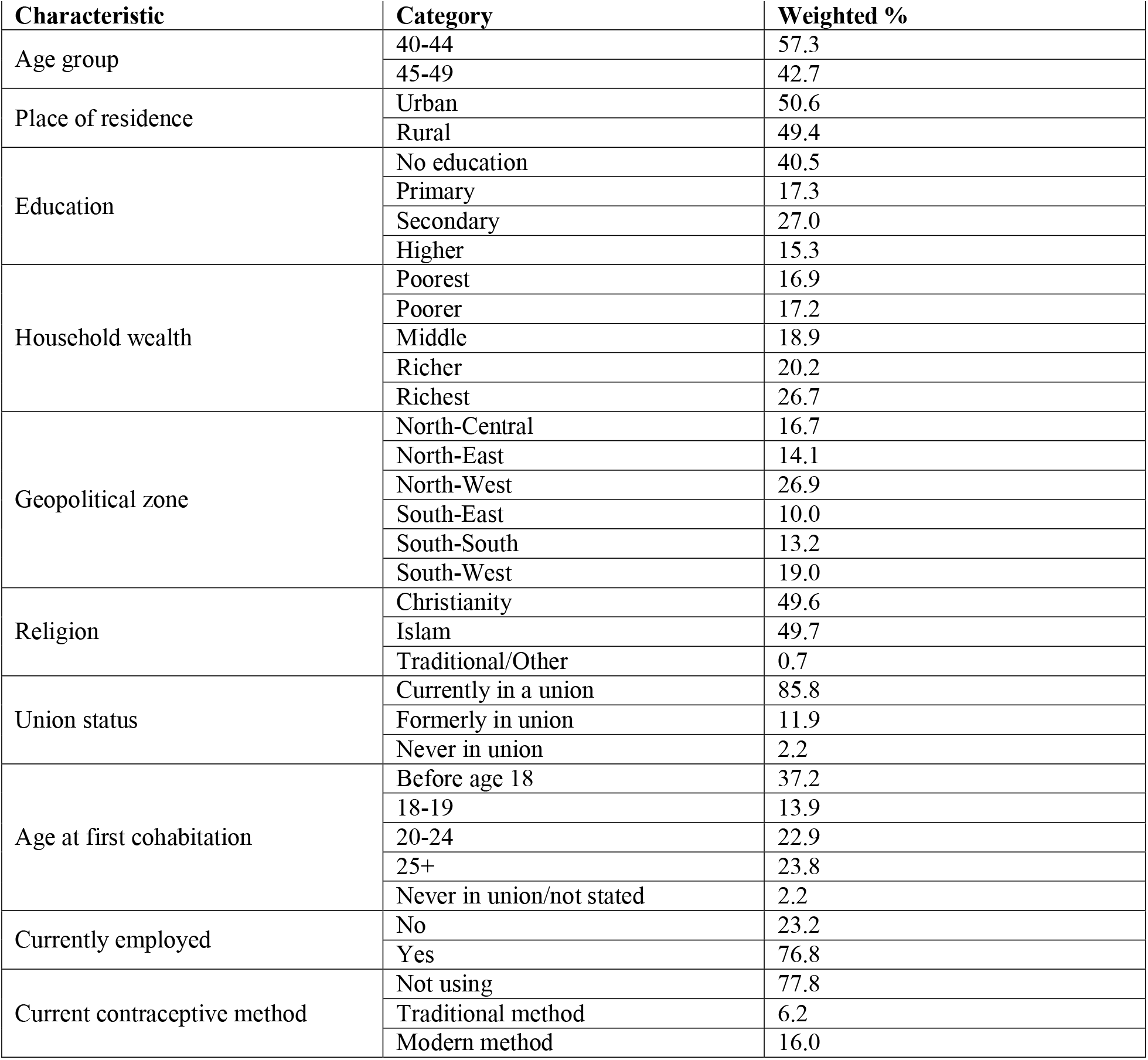
Weighted profile of women aged 40-49, 2024 Nigeria Demographic and Health Survey.

| Characteristic | Category | Weighted % |
| --- | --- | --- |
| Age group | 40-44 | 57.3 |
|  | 45-49 | 42.7 |
| Place of residence | Urban | 50.6 |
|  | Rural | 49.4 |
| Education | No education | 40.5 |
|  | Primary | 17.3 |
|  | Secondary | 27.0 |
|  | Higher | 15.3 |
| Household wealth | Poorest | 16.9 |
|  | Poorer | 17.2 |
|  | Middle | 18.9 |
|  | Richer | 20.2 |
|  | Richest | 26.7 |
| Geopolitical zone | North-Central | 16.7 |
|  | North-East | 14.1 |
|  | North-West | 26.9 |
|  | South-East | 10.0 |
|  | South-South | 13.2 |
|  | South-West | 19.0 |
| Religion | Christianity | 49.6 |
|  | Islam | 49.7 |
|  | Traditional/Other | 0.7 |
| Union status | Currently in a union | 85.8 |
|  | Formerly in union | 11.9 |
|  | Never in union | 2.2 |
| Age at first cohabitation | Before age 18 | 37.2 |
|  | 18-19 | 13.9 |
|  | 20-24 | 22.9 |
|  | 25+ | 23.8 |
|  | Never in union/not stated | 2.2 |
| Currently employed | No | 23.2 |
|  | Yes | 76.8 |
| Current contraceptive method | Not using | 77.8 |
|  | Traditional method | 6.2 |
|  | Modern method | 16.0 |

Geographically, the largest proportion of women resided in the North-West (26.9%), followed by the South-West (19.0%) and North-Central (16.7%). Women from the South-East constituted the smallest proportion of the sample (10.0%). Religious affiliation was almost evenly divided between Islam (49.7%) and Christianity (49.6%), while less than 1% practised traditional or other religions. Most women were currently in a marital or cohabiting union (85.8%), whereas 11.9% were formerly in a union and 2.2% had never been in a union. Early entry into marital or cohabiting unions was common, with 37.2% first cohabiting before age 18 and a further 13.9% doing so between ages 18 and 19. In contrast, 23.8% first cohabited at age 25 or later.

More than three-quarters of the women were employed (76.8%), while 23.2% were not. Despite their advanced reproductive ages, contraceptive use remained relatively limited. More than three-quarters were not using any contraceptive method (77.8%), while 16.0% were using a modern method and 6.2% were using a traditional method. Overall, the study population was characterized by substantial educational disadvantage, high levels of current union and employment, frequent early entry into union, and low contraceptive use.

### Fertility Differentials by Social and Demographic Characteristics

Table 2 presents variations in near-completed fertility among women aged 40–49 years by selected characteristics. Overall, women in this age group had an average of 5.53 children ever born, while 59.6% had given birth to five or more children. Fertility was substantially higher among rural women, who had an average of 6.39 children, compared with 4.68 children among urban women. Correspondingly, nearly three-quarters of rural women had five or more children (73.7%), compared with 45.8% of urban women. Women aged 45–49 years reported slightly higher fertility than those aged 40–44 years, with mean numbers of children ever born of 5.77 and 5.34, respectively.

**Table 2.**
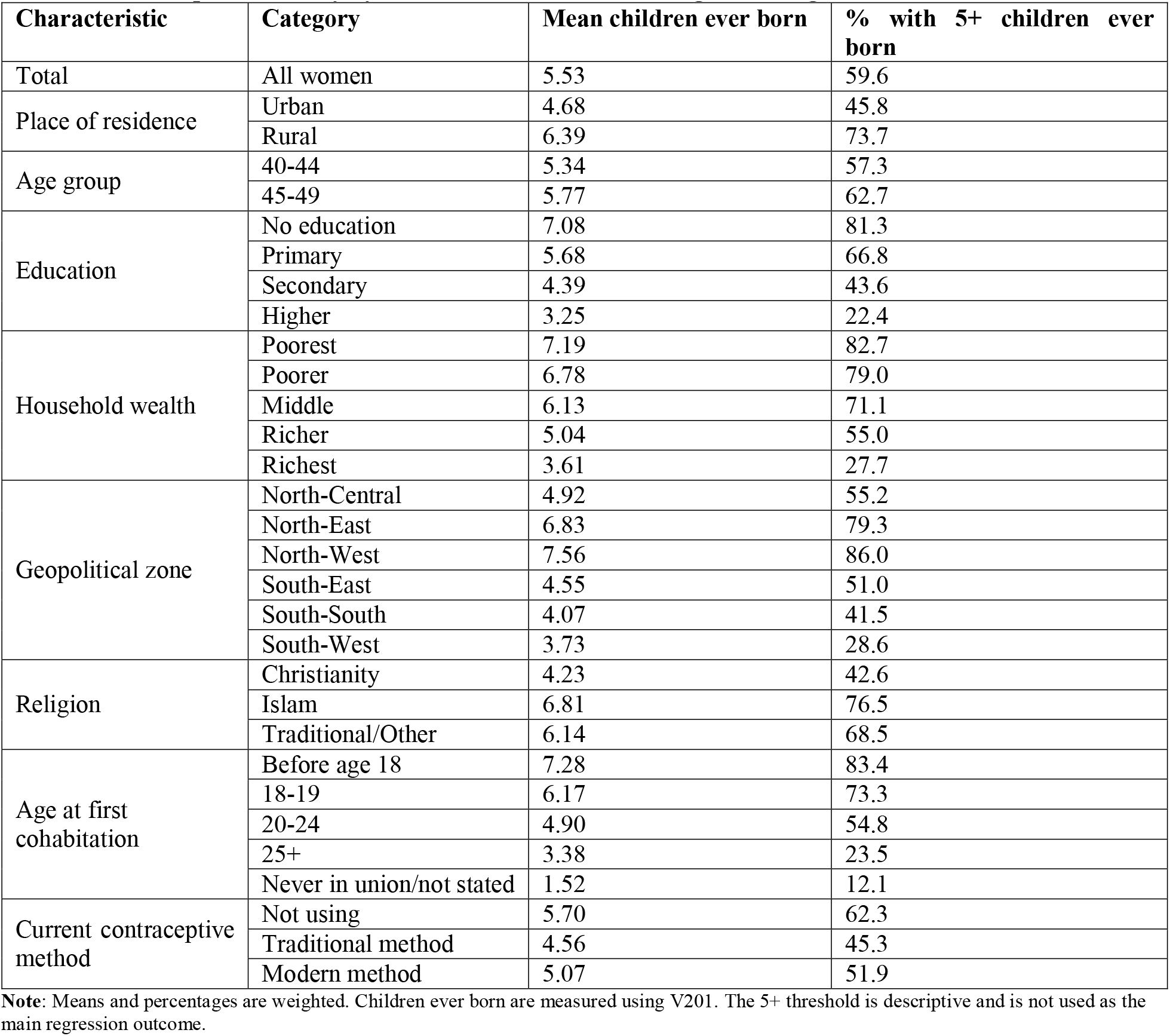
Near-completed fertility by selected characteristics among women aged 40-49.

Near-completed fertility declined consistently with increasing educational attainment. Women with no formal education had an average of 7.08 children, and 81.3% had five or more children. This decreased to 5.68 children among women with primary education, 4.39 among those with secondary education, and 3.25 among women with higher education. Only 22.4% of women with higher education had five or more children, compared with more than four-fifths of women with no education. A similar gradient was observed across household wealth groups. Women in the poorest households had an average of 7.19 children, and 82.7% had 5 or more, whereas women in the richest households had an average of 3.61 children, with 27.7% having 5 or more.

Distinct regional differences were also observed. Women in the North-West had the highest average number of children ever born at 7.56, followed by those in the North-East at 6.83. The percentage with five or more children was also highest in these regions, 86.0% in the North-West and 79.3% in the North-East. Conversely, women in the South-West had the lowest average fertility, at 3.73 children, and only 28.6% had five or more children. The South-South and South-East regions also showed relatively low fertility rates of 4.07 and 4.55, respectively. Muslim women averaged 6.81 children, compared to 4.23 among Christian women, while women of traditional or other religions averaged 6.14 children. Over 75% of Muslim women had five or more children, in contrast to 42.6% of Christian women.

Fertility also varied considerably by age at first cohabitation. Women who entered a union before age 18 had the highest mean number of children ever born (7.28), and 83.4% had five or more children. Fertility declined progressively as age at first cohabitation increased, reaching an average of 3.38 children among women who first cohabited at age 25 or later. Only 23.5% of this group had five or more children. Women who had never entered a union or whose age at first cohabitation was not stated had the lowest mean fertility of 1.52 children. Regarding contraceptive use, women who were not currently using contraception had the highest mean number of children ever born at 5.70, and the largest proportion with five or more children at 62.3%. Overall, the descriptive results show that higher near-completed fertility was concentrated among rural women, those with limited education, women from poorer households, residents of northern geopolitical zones, Muslim women, and those who entered unions at younger ages.

### Residence-Specific Composition of Key Mechanisms

Table 3 presents the residence-specific composition of selected educational, socioeconomic, and union-formation characteristics among women aged 40–49 years. Clear urban–rural differences were evident across all three mechanisms. Urban women were substantially more educated than rural women. More than three-fifths of urban women had attained at least secondary education (60.8%), comprising 37.1% with secondary education and 23.7% with higher education. In contrast, 60.0% of rural women had no formal education, while only 23.3% had attained secondary or higher education. Primary education accounted for a similar proportion of women in urban and rural areas, at 17.8% and 16.7%, respectively.

**Table 3:** Residence-specific composition of key mechanisms.

| Characteristic | Urban weighted % | Rural weighted % |
| --- | --- | --- |
| **Education** |  |  |
| No education | 21.4 | 60.0 |
| Primary | 17.8 | 16.7 |
| Secondary | 37.1 | 16.7 |
| Higher | 23.7 | 6.6 |
| **Wealth** |  |  |
| Poorest | 3.3 | 30.9 |
| Poorer | 6.3 | 28.4 |
| Middle | 15.5 | 22.4 |
| Richer | 29.2 | 11.0 |
| Richest | 45.7 | 7.3 |
| **First cohabitation** |  |  |
| Before age 18 | 25.6 | 49.0 |
| 18-19 | 12.8 | 15.1 |
| 20-24 | 26.3 | 19.4 |
| 25+ | 32.3 | 15.0 |
| Never in union/not stated | 3.0 | 1.4 |

**Note.** Percentages are column percentages within residence categories. The table shows only the main compositional mechanisms emphasized in the analysis.

Household wealth was also strongly differentiated by residence. Nearly three-quarters of urban women belonged to the richer or richest wealth categories (74.9%), including 45.7% in the richest quintile. By comparison, 59.3% of rural women were concentrated in the poorest or poorer wealth categories, while only 18.3% were in the richer or richest wealth categories. The proportion in the poorest wealth quintile was almost ten times higher among rural women than among urban women, at 30.9% and 3.3%, respectively.

Residence differences were similarly apparent in the timing of first cohabitation. Almost half of rural women first entered a union before age 18 (49.0%), compared with approximately one-quarter of urban women (25.6%). When women who first cohabited at ages 18–19 were included, 64.1% of rural women had entered a union before age 20, compared with 38.4% of urban women. Conversely, 58.6% of urban women first cohabited at age 20 or later, compared with 34.4% of rural women. Overall, the table shows that rural residence was characterized by lower educational attainment, greater concentration in poorer households, and earlier union entry. In contrast, urban residence was associated with higher education, greater household wealth, and delayed cohabitation.

### Sequential Count Models

Table 4 presents four sequential survey-weighted Poisson regression models predicting the number of children ever born among women aged 40-49. Incidence rate ratios are interpreted relative to urban residence, age 40-44 years, no education, the poorest wealth quintile, first cohabitation before age 18, Christianity, North-Central residence, not currently employed, and no current contraceptive use.

**Table 4.**
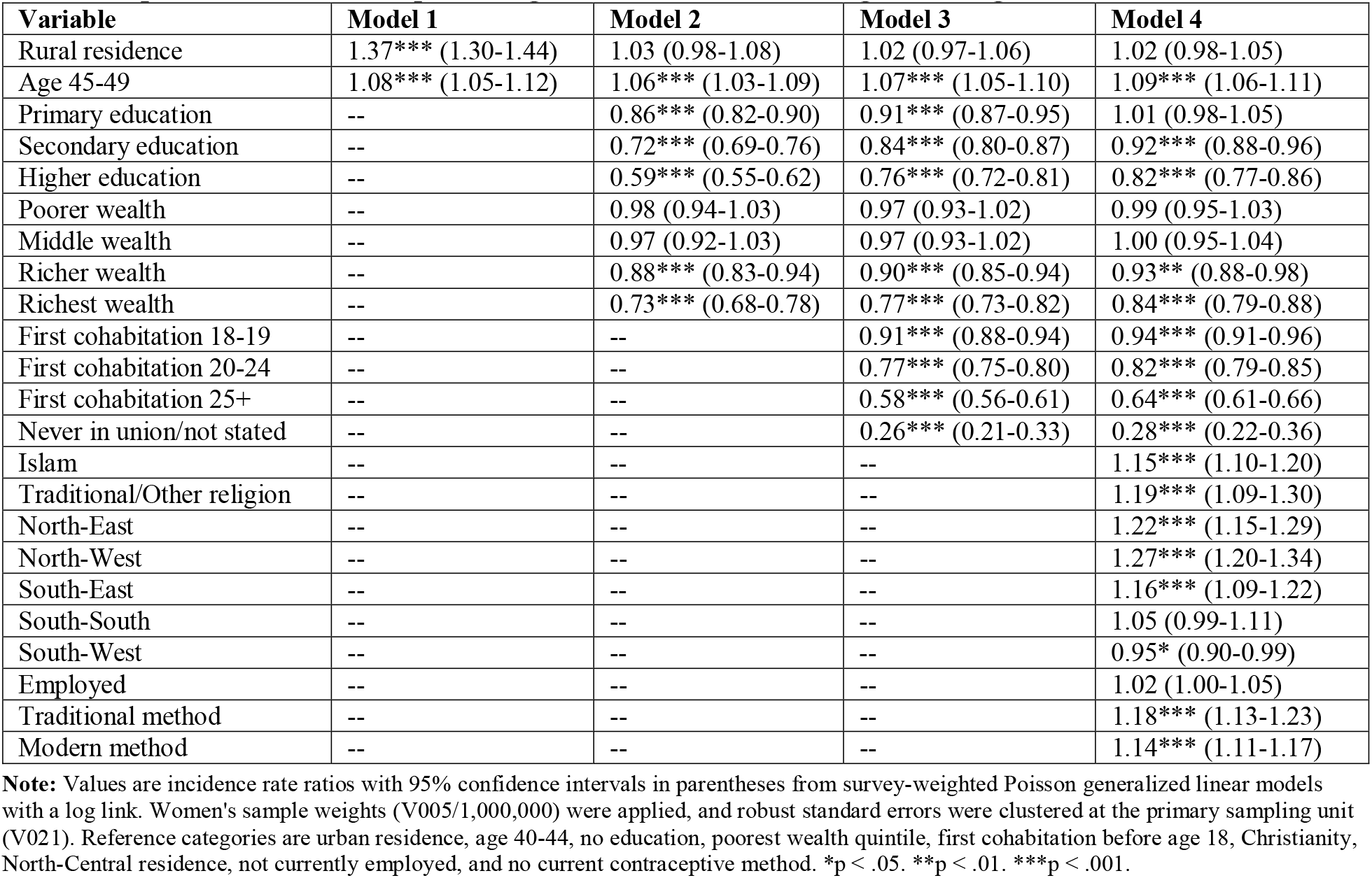
Sequential count models predicting children ever born among women aged 40-49.

In Model 1, rural residence was positively and significantly associated with the number of children ever born. Rural women had an expected number of children that was 37% higher than that of urban women (IRR = 1.37, p < .001). However, after education and household wealth were introduced in Model 2, the magnitude of the association declined to 3% and was no longer statistically significant. Rural residence remained statistically insignificant in Models 3 and 4, indicating that the initial rural–urban fertility difference was largely explained by differences in education, household wealth, age at first cohabitation, and other characteristics included in the models. Women aged 45–49 consistently had significantly more children than those aged 40–44. In the fully adjusted model, women aged 45–49 had an expected number of children that was 9% higher than that of women aged 40–44 (IRR = 1.09, p < .001).

Educational attainment was inversely associated with the number of children ever born. In Model 2, compared with women with no education, those with primary, secondary, and higher education had 14%, 28%, and 41% fewer children, respectively, and all associations were statistically significant (p < .001). The magnitudes of these associations decreased after age at first cohabitation was introduced in Model 3. In the fully adjusted model, primary education was no longer significantly associated with the number of children ever born. However, women with secondary education had 8% fewer children (IRR = 0.92, p < .001), and those with higher education had 18% fewer children (IRR = 0.82, p < .001) than women with no education.

Household wealth also showed an inverse association with fertility, though the pattern was concentrated in the upper wealth categories. Compared with women in the poorest households, those in the poorer and middle wealth categories did not have significantly different numbers of children across any of the models. In the fully adjusted model, women in richer households had 7% fewer children (IRR = 0.93, p < .01). In comparison, those in the richest households had 16% fewer children (IRR = 0.84, p < .001) than women in the poorest households.

Age at first cohabitation was strongly and negatively associated with the number of children ever born, and compared with women who first cohabited before age 18, those who first cohabited at ages 18–19 had 6% fewer children (IRR = 0.94, p < .001) in the fully adjusted model. Those who first cohabited at ages 20–24 had 18% fewer children (IRR = 0.82, p < .001), while women who first cohabited at age 25 or later had 36% fewer children (IRR = 0.64, p < .001). Women who had never entered a union or whose age at first cohabitation was not stated had 72% fewer children than those who first cohabited before age 18 (IRR = 0.28, p < .001). The progressively lower incidence rate ratios indicate that later union entry was associated with substantially lower near-completed fertility.

After adjusting for other characteristics, religion remained significantly associated with the number of children ever born. Compared with Christian women, Muslim women had 15% more children (IRR = 1.15, p < .001), and women affiliated with traditional or other religions had 19% more children (IRR = 1.19, p < .001).

Significant regional differences were also observed. Relative to women in North-Central Nigeria, women in the North-East had 22% more children (IRR = 1.22, p < .001), those in the North-West had 27% more children (IRR = 1.27, p < .001), and those in the South-East had 16% more children (IRR = 1.16, p < .001). Women in the South-West had 5% fewer children than those in North-Central Nigeria (IRR = 0.95, p < .05). Women in the South-South had 5% more children than those in North-Central Nigeria, but the difference was not statistically significant.

Current employment was positively associated with the number of children ever born, but the association was not significant. Employed women had an expected number of children 2% higher than that of non-employed women, but the difference was not statistically significant. Current contraceptive use was positively and significantly associated with children ever born. Compared with women who were not using contraception, women using traditional methods had 18% more children (IRR = 1.18, p < .001), while modern-method users had 14% more children (IRR = 1.14, p < .001). Overall, the fully adjusted model indicates that age, secondary and higher education, upper household wealth, age at first cohabitation, religion, geopolitical zone, and current contraceptive use were significantly associated with near-completed fertility. In contrast, residence and employment were not independently associated with the number of children ever born.

### Synthesis of Results

Taken together, the results support a compositional interpretation of the rural-urban fertility gap. Rural women had markedly higher near-completed fertility in the descriptive and age-adjusted analyses. However, the residence coefficient was substantially attenuated after education and household wealth were included and remained nonsignificant after union timing and the remaining covariates were added. The observed gap, therefore, reflects the spatial concentration of educational disadvantage, economic deprivation, early union formation, and regional differences rather than an independent effect of residence alone.

## Discussion and Conclusion

### Principal Findings

This study explored rural-urban differences in nearly completed fertility among Nigerian women aged 40-49 using data from the 2024 NDHS. Three key findings emerged. First, there was a substantial rural-urban gap: rural women had an average of 6.39 children ever born, compared to 4.68 for urban women, with nearly 75% of rural women having at least five children. Second, the initially significant rural coefficient was almost entirely nullified after accounting for education and household wealth. Residence was no longer significant after controlling for age at first cohabitation, religion, geopolitical zone, employment, and contraceptive use. Third, factors such as later union entry, higher education levels, greater household wealth, and regional context remained strongly linked to cumulative fertility in the fully adjusted model.

The significant reduction in the residence coefficient indicates that the main difference between rural and urban areas lies in their composition. Rural women face more social conditions linked to higher cumulative childbearing. As shown in Table 3, 60.0% of rural women lacked formal education, 59.3% belonged to the poorest or poorer wealth quintiles, and 49.0% started cohabiting before age 18. In contrast, urban women had figures of 21.4%, 9.6%, and 25.6%, respectively. Therefore, rural residence functions more as a spatial grouping of unequal life chances than as a standalone demographic factor. This perspective avoids attributing rural fertility solely to culture or preference and instead ties it to broader institutional factors, such as education, material security, and the timing of family formation.

Education remains linked to lower fertility, but the effect diminishes after accounting for social factors. Women with secondary education have 8% fewer children, while those with higher education have 18% fewer than women with no education. The disappearance of the primary-education effect in Model 4 suggests that limited schooling alone may not significantly affect total fertility when factors such as union timing, religion, region, and social context are taken into account. This aligns with evidence that the fertility-reducing benefits of education are more pronounced at secondary and higher levels, where schooling often delays marriage, broadens employment goals, enhances reproductive knowledge, and boosts women’s bargaining power (Ainsworth et al., 1996; Kravdal, 2002; Liu & Raftery, 2020; Olowolafe et al., 2025).

The concentration of wealth was mainly in the top quintiles. After adjustments, women from poorer and middle households showed no significant difference from those in the poorest group, while women from wealthier households tended to have fewer children. This indicates a threshold effect rather than a simple linear trend. Material benefits may need to reach a certain level to significantly influence factors like schooling, housing, child-rearing costs, transportation, access to services, and expectations for investing in each child. Caldwell’s wealth-flows theory remains relevant: fertility tends to decline as the economic and social advantages shift from having many children to investing more in each child (Caldwell, 1976, 1982).

Age at first cohabitation emerged as the most evident life-course factor. Compared to women who entered a union before 18, those who first cohabited at 18-19, 20-24, and 25 or older tended to have fewer children. This aligns with the proximate-determinants framework, as later union formation reduces exposure to marital childbearing and allows more time for schooling, employment, and the development of reproductive autonomy (Bongaarts, 1978; Casterline et al., 2017). This pattern should not be viewed solely as a behavioral choice, since early cohabitation is socially influenced by factors such as educational opportunities, household poverty, family expectations, and local marriage customs.

Regional differences in Nigeria indicate that there is no single fertility regime across the country. Women in the North-East and North-West regions experienced significantly higher expected cumulative fertility compared to those in North-Central Nigeria. Meanwhile, women in the South-West had slightly lower fertility levels. These regional disparities remained even after accounting for factors such as education, wealth, union timing, religion, employment, and contraceptive use. This suggests that broader contextual factors, such as marriage practices, gender relations, school participation, conflict and insecurity, child health environments, infrastructure, and family-size preferences, also play a role. Prior research on Nigeria also highlights considerable regional variation in fertility timing and levels (NPC & ICF, 2019; Olowolafe et al., 2023, 2025).

The positive link between current contraceptive use and the total number of children ever born should be understood through a life-course perspective. For women aged 40-49, the current method used often happens after they have reached high parity, decided to stop having children, or want to avoid health risks associated with later pregnancies. Thus, current use does not reflect overall contraceptive exposure throughout their reproductive life. This correlation could result from reverse timing or a tendency for women with many children to adopt contraceptive methods; it should not be seen as evidence that contraception increases fertility. When studying fertility in older women, it is important to distinguish behaviors such as initiating, spacing, discontinuing, switching, and stopping contraception, rather than viewing method use at the time of interview as a measure of lifetime exposure (Rossier & Corker, 2017; Tsui et al., 2017).

### Contribution to the Field

The study advances fertility research in four key ways. Firstly, it uses the latest nationally representative NDHS data to examine cumulative childbearing among women near the end of their reproductive years. While most national analyses focus on period fertility, recent births, or all women aged 15-49, centering on women aged 40-49 offers a near-cohort view on the reproductive impacts of conditions experienced throughout adolescence and adulthood.

Second, the sequential modeling approach separates the observed rural-urban gap from the social makeup of rural and urban populations. The residence coefficient no longer appears once education and wealth are included, showing that the unequal distribution of resources and opportunities across the life course can statistically explain place-based differences. This provides a clearer interpretation than explanations that treat rural residence as a static cultural factor.

Third, the analysis highlights age at first cohabitation as a key factor linking structural inequality to near-complete fertility. Education and wealth influence this process by affecting when women start cohabiting and how long they remain subject to socially approved childbearing. Fourth, the study explains the importance of using both period and cumulative fertility indicators. Period measures reflect current fertility conditions, while the number of children ever born among women aged 40-49 shows the long-term effects of earlier education, marriage, reproductive health, and household environments.

### Policy Implications

The policy implication is clear: narrowing rural fertility disparities requires more than expanding the contraceptive supply. Family planning services remain essential, but the residence gap stems from upstream factors in education, material resources, and union timing. Policies should therefore combine reproductive health provision with sustained investment in girls’ secondary education, school completion, safe transportation to school, protection against school exclusion following pregnancy or marriage, and credible enforcement of the legal minimum age at marriage.

Interventions should be life-course specific. During adolescence, priorities include school retention, prevention of child marriage, comprehensive sexuality education, and delayed first birth. During early and middle adulthood, programs should emphasize birth spacing, method choice, continuity of care, and partner communication. For women approaching the end of reproductive life, services should support safe stopping, management of contraindications, and prevention of unintended later-age pregnancies. A uniform family planning message cannot address these different reproductive stages.

Regional targeting remains essential. The ongoing fertility differences in the North-East and North-West show that relying on national averages and uniform programs will not address key local constraints. Strategies should be developed collaboratively with women’s groups, schools, primary health-care providers, religious authorities, and traditional leaders, while ensuring women’s reproductive rights are protected and avoiding coercive population-control methods. Evaluation of programs should consider not only contraceptive use but also metrics like school completion, age at union and first birth, birth spacing, unmet need, and women’s capacity for informed reproductive choices.

Since the outcome reflects births accumulated over many decades, the results also clarify when policy effects are likely to appear. Investments today in girls aged 10-19 will not immediately affect the fertility of women aged 40-49, who are nearly finished with their fertility. Instead, their main demographic impacts will become visible in younger groups as education, union formation, and reproductive paths evolve. Therefore, policy evaluation should use both short-term indicators and cohort-sensitive measures that can detect these delayed structural effects.

### Limitations and Future Research

Several limitations should be considered when interpreting these findings. The NDHS data is cross-sectional, which means it cannot establish cause-and-effect relationships or temporal sequences. Changes in residence over time, due to migration, mean that current residence might differ from that during earlier childbirth periods. Household wealth and employment data were collected at the interview point, not throughout the entire reproductive lifespan. Consequently, the observed reduction in the residence coefficient likely reflects compositional factors rather than a formal mediation analysis. The “children ever born” metric is cumulative and lacks details on the timing, spacing, intent, or partnership circumstances of births. It is also susceptible to recall bias, though birth histories tend to be more accurate for living and recent births. Limiting the sample to women aged 40-49 approximates completed fertility, but some women aged 40-44 may still have additional children. The analysis excludes women who survived to the survey but were not interviewed, which could introduce bias due to differences in mortality or response rates. Additionally, current contraceptive use may be subject to reverse causality, as it could be a consequence of, rather than a predictor of, high parity.

The models did not include full contraceptive histories, fertility preferences over time, husband or partner characteristics, child mortality, media exposure, local service quality, community gender norms, or measures of migration history. Sampling weights and primary-sampling-unit-clustered robust standard errors were used. However, future analyses should replicate the findings using full design-based variance estimation that explicitly incorporates strata and multilevel models that separate individual, community, state, and regional effects.

Future research should explore three key areas. First, parity-progression and event-history models should analyze the transition to multiple births, birth intervals, and stopping behaviors. Second, decomposition methods should estimate the proportion of the rural-urban gap caused by differences in characteristics versus differences in coefficients. Third, data from multiple NDHS rounds should be used to compare successive cohorts of women aged 40-49, assessing whether the residence gap is shrinking and identifying if this change is due to educational expansion, delayed union formation, shifts in wealth, or altered fertility responses to these factors.

## Conclusion

Near-completed fertility remains markedly higher among rural than urban Nigerian women aged 40-49. However, multivariable results show that residence is not independently associated with cumulative births after controlling for education, wealth, union timing, and broader social context. The national rural-urban difference is chiefly the demographic expression of unequal life-course conditions. Women with secondary or higher education, those in the upper wealth quintiles, and those who entered unions later had substantially fewer children, while important regional differences persisted. A durable reduction in fertility inequality will therefore depend on policies that alter the conditions under which reproductive careers begin and unfold. Keeping girls in secondary school, delaying early unions, improving material security, and ensuring accessible and acceptable reproductive health services are not separate policy agendas; they are interconnected components of the fertility transition. Framing rural fertility as a structural and life-course issue provides a more accurate diagnosis and a more actionable policy response than attributing the gap solely to residence or culture.

## Data Availability

The 2024 Nigeria Demographic and Health Survey datasets are available to registered researchers through The DHS Program data portal, subject to approval of a research request and compliance with the data-use agreement. The present analysis used a woman's individual-recode extract restricted to ages 40-49. Access was requested on May 3, 2026, and authorized by ICF on May 4, 2026.

